# Experiences of Healthcare Providers on Labor and Delivery with Values Clarification Workshops Prior to Implementing Induction Abortion Services

**DOI:** 10.64898/2026.01.23.26344558

**Authors:** Laura Jacques, Elise Cowley, Laurie Lapp, Jacquelyn Askins, Anna Altshuler

**Affiliations:** Department of Obstetrics and Gynecology, University of Wisconsin-Madison; University of Wisconsin-Madison School of Medicine and Public Health; Alameda Health Systems

## Abstract

**Objective:** To evaluate experiences of labor and delivery (L&D) staff with a Values Clarification and Attitude Transformation (VCAT) workshop on abortion perspectives and professional responsibilities prior to expanding abortion services to L&D.

**Methods:** We conducted VCAT workshops about abortion with 192 perinatal healthcare personnel at a single urban, community-based, tertiary care center in California. The workshops used a virtual adaptation of the *Four Corners* exercise, a previously published VCAT method that prompts participants to reflect on and represent diverse abortion viewpoints. Post-workshop, 25 participants, including nurses, physicians and midwives completed semi-structured interviews about their experience with the workshop. Transcripts were coded and thematically analyzed using a framework approach.

**Results:** Four major themes emerged: *(1) VCAT promoted personal reflection and perspective taking*, where participants described how representing others’ views deepened their understanding of their own beliefs, fostered empathy, and challenged assumptions about colleagues. *2) VCAT prompted reconsideration of professional roles and the role of L&D in abortion care* with some viewing L&D as a space incompatible with abortion care and others seeing opportunity to expand inclusive, compassionate care. *(3) Professional and personal contexts shaped participants’ engagement with the workshop* with factors like compassion fatigue from many years of service in a hospital with many social needs and personal abortion experiences influencing how participants processed the workshop. *(4) Respect for autonomy and commitment to patient care emerged as core, shared values*, even among those with differing personal beliefs.

**Conclusions:** VCAT workshops provided a structured opportunity for L&D staff to reflect on the complex personal and professional dimensions of abortion care. The process helped participants explore value tensions, clarify their professional roles, and identify shared ethical commitments to patient care. These findings suggest VCAT may be a valuable tool for preparing multidisciplinary perinatal teams to navigate institutional abortion care expansion with empathy, professionalism and cohesion.

## INTRODUCTION

Pregnant people facing complications in the second trimester should have the option to end their pregnancy in a safe manner that aligns with their medical needs and preferences.^1,2^ Medication-induced abortion is comparable in safety and effectiveness to contemporary surgical method, and national and international organizations support access to both approaches.^3–6^ Despite this broad organizational support, most patients in the United States who are able to access second-trimester abortion receive surgical care.^7^ The predominance of surgical over medical management reflects the historical safety advantages of these techniques, as well as the longstanding concentration of abortion services within dedicated facilities that primarily offer surgical services^.8^ As a result, current patterns of care may reflect structural and historical constraints rather than patient preference and may not represent the choices patients would make if both options were routinely available.

While not traditionally viewed as sites for abortion care, hospital-based Labor & Delivery (L&D) units possess substantial clinical expertise to provide later second-trimester abortion services via labor induction. Moreover, in the current political climate—characterized by reduced availability of abortion providers and rising demand for services—expanding abortion care within hospital-based L&D units may represent a practical and necessary strategy to improve access.^9–11^

Barriers to providing abortion care within L&D settings are multifactorial and include legal, institutional, and individual-level constraints. Even in states with fewer legal restrictions, hospitals or specific units may implement internal policies that limit service provision or lack a sufficiently prepared and willing workforce. L&D teams already possess the technical skills required to provide medication-induced abortion, as the clinical process is identical to labor induction for pregnancy loss or previable premature rupture of membranes in the second trimester. What distinguishes abortion care is not clinical complexity, but stigma.^12^ At a systems level, stigma influences resource allocation, including staffing, physical environments that enable abortion care, standard policies and procedures, and staff support services.^13,14^ At the individual level, stigma can affect professional reputation and clinicians’ sense of belonging within their care teams.^15,16^

For teams seeking workforce preparedness, there is currently no clear or standardized pathway for becoming trained and supported to provide abortion services within hospital-based L&D units. The Values Clarification and Attitude Transformation (VCAT) workshop has been widely described as an intervention to address psychosocial barriers to abortion provision among healthcare workers).^17–21^ VCAT is designed to help health professionals reflect on their personal values related to abortion and to think about those values in the context of their professional roles and responsibilities, rights, and limits of conscientious objection.^19^ To inform workforce preparedness in this stigmatized and operationally complex setting, we explored L&D clinicians’ experiences with a VCAT workshop delivered in advance of implementing induction abortion services on the unit.

## METHODS

### Study Setting and Participants

This study took place at a large, urban, publicly funded safety net hospital in Northern California serving a racially and socioeconomically diverse patient population. The labor and delivery (L&D) unit is staffed by hospital-employed Obstetrician Gynecologists (OB-GYNs) and midwives, averages 1,200 deliveries per year, and operates a Level II neonatal intensive care unit (NICU). An interdisciplinary team of nursing, physician and midwifery leaders representing Obstetrics, Midwifery & Gynecology; Pediatrics; and Pathology and Laboratory Medicine departments came together to devise and implement induction abortion services on their L&D Unit. As the team identified personnel willingness—shaped by stigma surrounding abortion, particularly second-trimester abortion—as the primary barrier to implementation in a setting oriented toward optimizing outcomes for birthing people and their babies, preparedness efforts centered on addressing this barrier as a matter of equity, ensuring that institutional values and clinical practices aligned to support all patients’ reproductive health needs. Prior to VCAT workshops, every staff member was required to attend a 30-minute interactive presentation explaining the rationale for launching this service on Labor & Delivery.

### Workshop Description

All perinatal care employees were required to attend one workshop as part of the institutional rollout of induction abortion services. Workshops were offered at time aligned with both day and night shifts. Participants could choose to participate in the workshops either in-person or virtually and final sessions were held around people’s preferences. Workshops were modeled after “Four Corners^”22,23^ a structured reflective exercise designed to prompt participants to consider diverse perspectives on abortion. Workshops were intentionally stratified by professional role, with separate sessions for providers (OB-GYNs and midwives) and for nurses and staff (L&D, NICU, postpartum nurses and other staff. Each session was facilitated virtually by one of three trained VCAT facilitators external to the study site.).

### Participant Recruitment and Interviews

Participants were invited to voluntary interview via email and offered a $100 Amazon Gift Card as remuneration. One study member (JA) conducted all 45–60-minute interviews via Zoom from November 06, 2023 to December 20, 2023. We aimed to include a range of clinical roles and ceased recruitment upon reaching thematic saturation. Interviews were audio-recorded, transcribed verbatim, and deidentified using participant identification numbers that were not known to anyone outside of the research study staff.

Our research team consisted of two OBGYNs and qualitative researchers (LJ, AA), a departmental communications director with a background in journalism (JA), and an MD PhD (EC) an MD MPH (LL) student.

### Analysis and Ethical Review

We analyzed interview transcripts using a combined inductive-deductive approach in NVivo. Two researchers (EC, LL) independently coded the data and iteratively refined a codebook for clarity and consistency. They resolved discrepancies through discussion until reaching consensus. Using a framework approach^24^, we identified and categorized key themes and subthemes. To ensure rigor, we documented coding decisions, engaged in peer debriefing, reviewed disconfirming evidence and refined final themes through team discussion.

This study was reviewed and approved by the Alameda Health System Institutional Review Committee (IRB #2023-0349). The University of Wisconsin-Madison Institutional Review Board determined the project to be exempt as a quality improvement initiative. A Data Use Agreement was established between Alameda Health Systems and the University of Wisconsin to support secure data sharing and ensure compliance with institutional privacy requirements.

## RESULTS

We had 192 healthcare workers (155 nurses and L&D staff, 19 midwives, and 18 OBGYNs) participate in six workshops between August and September 2023. All perinatal service providers were required to attend one VCAT workshop as a condition of employment during institutional abortion service implementation. Twenty-five participants (8 midwives, 1 OB-GYN, 15 nurses, and 1 L&D staff) volunteered for interviews post-workshop (Figure 1). Interviewees had a mean of 17.5 years (range 0.5-43 years) in practice and 7.5 years (range 0.5-39 years) at the institution.

**Figure 1.**
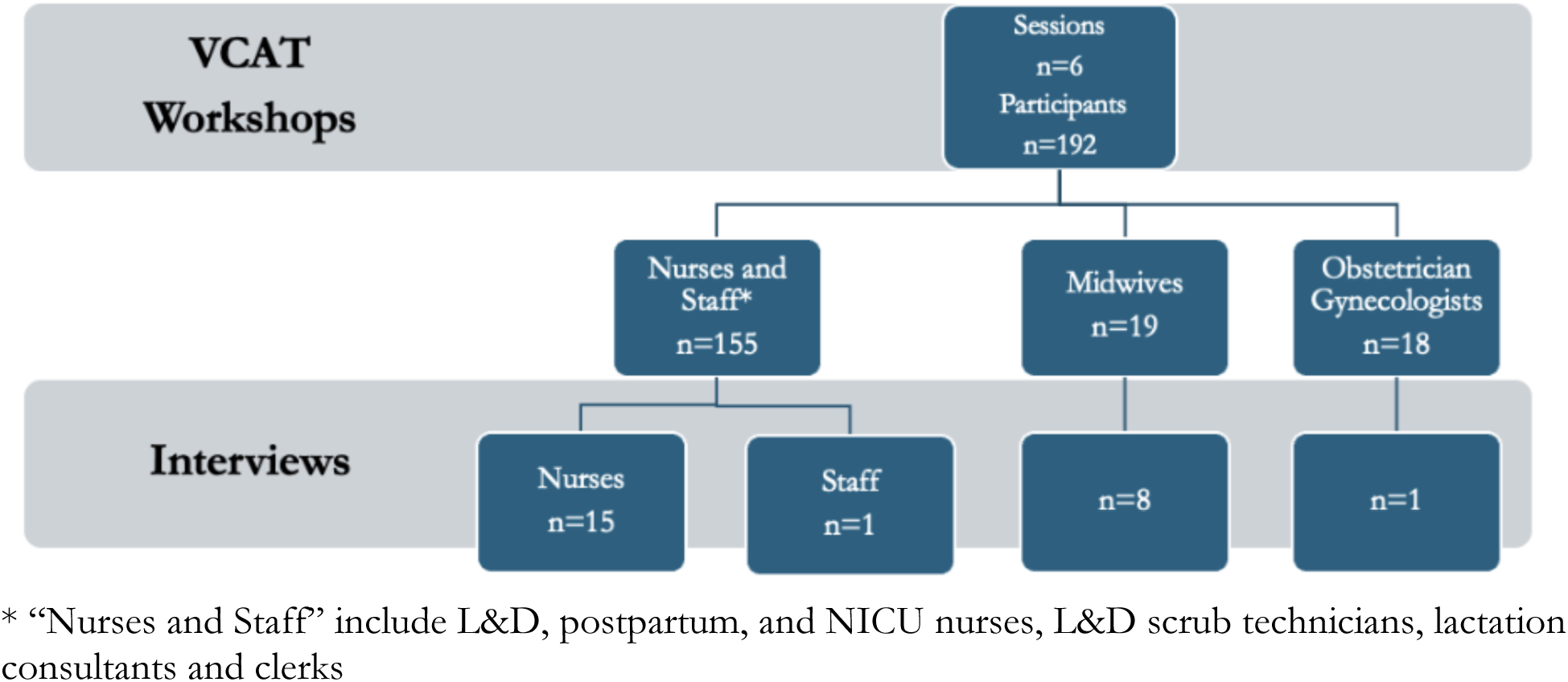
Consort Diagram of VCAT Workshop and Interview Participants

We identified four major themes in perinatal care providers’ experiences with the VCAT workshop: (1) VCAT promoted personal reflection and perspective taking, 2) VCAT prompted reconsideration of professional roles and the role of L&D in abortion care, (3) Professional and personal contexts shaped participants’ engagement with the workshop, (4) Respect for autonomy and commitment to patient care emerged as core, shared values.

### 1. VCAT promoted personal reflection and perspective taking

While most participants’ core beliefs about abortion did not change, the workshop helped clarify views and provided a framework for respectful discussion with colleagues. A midwife described the process of personal reflection: *“I walked out of the workshop feeling more relaxed and confident that the work I’m doing is ethical for me and consistent with my morality”* (1012, midwife). In addition to prompting reflection on one’s own ethics, a core component of VCAT is encouraging consideration of others’ perspectives. Analysis of participants’ reflections on engaging with colleagues’ ethical frameworks revealed experiences consistent with well-described components of clinical empathy building. We organized these descriptions of perspective-taking into subthemes using Hojat’s framework of clinical empathy^25^: *a) cognition and understanding, (b) communication of understanding and (c) intention to help*.

#### (a) Cognition and Understanding

During the workshop, participants were asked to consider varied positions on abortion care. Several participants appreciated how the workshop required them to consider the “*gray* zones” in abortion beliefs, and not “*just the ‘support abortion’ vs ‘against abortion’ dichotomy.”* (1005, midwife). A physician likewise described a deepened appreciation for the complexity of others’ values, noting that her understanding felt “l*ess simplistic than before*” and *“more attuned to nuance”* (2007, MD). Some participants recognized that they had made inaccurate assumptions about colleagues’ beliefs. One midwife reflected:

> *“I went in assuming I could guess what other people were feeling, especially people I’d worked with for years. I was completely humbled by how wrong I was. People’s feelings are complex, and that was really interesting to see.”* (1001, midwife)

#### (b) Communication of Understanding

During VCAT, participants were asked not only to describe but to defend others’ abortion-related viewpoints, pushing them to give voice to values they did not necessarily share. One L&D nurse reflected that the exercise helped her “*step out of my own mindset and take on someone else’s thought process*” by considering how experiences such as trauma shape beliefs (6015, L&D RN). A midwife also felt that the workshop helped her be able to represent others’ viewpoints, statin*g, “It wasn’t hard for me to understand or represent where that person was coming from. It made me think, these are all valid ways to feel, and you are welcome to feel them.”* (1012, midwife)

For a smaller group, empathy extended beyond understanding to intention and action. One midwife described envisioning how the midwifery model of care could be applied to abortion services:

> *“I think that midwives have the potential to bring abortion services to a holistic, complete person service, that’s not just done behind closed doors, but is done with support. Just like with birth, “you’re going through something that’s a really powerful moment in your life, and you deserve support in that time.” I think that there’s something about the midwifery model of care that could also apply to abortion. Abortion’s a big decision and it has an impact, whatever impact that is, it’s a big decision, and it has an impact on your life, and I think that it should be almost honored in a different way. . . I think that the midwifery model of care has the potential to bring abortion out of the darkness a little bit and start providing people with more comprehensive support.”*(1014, midwife).

### 2. VCAT stimulated reconsideration of professional roles and the purpose of L&D in the context of abortion care

Required alongside the expansion of abortion services from the operating room to labor and delivery, the VCAT workshop prompted many participants to reflect on the appropriate scope of care on a labor and delivery unit. Some—particularly NICU nurses—felt abortion services were at odds with the unit’s mission and should occur elsewhere: *“As a NICU nurse, my duty or my job is to save the babies’ lives and not to support taking their lives*” (6019, NICU RN). One nurse noted that although staff *“cared for the patients well,”* providing abortion care on labor and delivery left many feeling distressed behind the scenes (3023, NICU RN). Another NICU nurse described how labor and delivery’s identity as a space of wellness and birth made abortion care feel disruptive:

*“Labor and delivery promotes birth—this idea of wellness and happy situations. When there’s a demise, the spirit of the unit changes. If it’s a late abortion and you’re the L&D nurse, it puts your beliefs into play. It’s hard to separate that, especially if you’re used to healthy babies going home—and now there’s no baby going home.”* (5033, NICU RN).

In contrast, other participants viewed labor and delivery as a natural setting for abortion care and felt their existing skills could be leveraged to provide compassionate support. Several labor and delivery nurses described appreciating the opportunity to support patients and their community through abortion care, with one nurse noting that the workshop gave her “*another way to serve women that could really affect their lives in a more compassionate way”* (8012, L&D RN).

### 3. Professional and personal contexts shaped participants’ engagement with the workshop

Personal and professional experiences influenced how participants processed the VCAT workshop and interpreted their roles in abortion care. These contexts ranged from the ongoing strain of working in a busy, under resourced, high-risk hospital setting, to the emotional weight of their own abortion experiences–particularly as they considered offering the service to others. Participants also reflected on the post-*Roe vs Wade* political and legislative climate and its potential impact on their patients. Within this theme, we identified three integrated, but distinct subthemes *(a) Professional compassion fatigue (b) Personal abortion experiences influenced beliefs, and (c) Post-Roe context heightened VCAT relevance*.

#### (a) Professional Compassion Fatigue

Participants described the emotional toll of labor and delivery work, shaped by a dichotomy between public expectations of joy and the clinical realities of grief and loss. Over time, the cumulative exposure to tragedy and unmet expectations can erode empathy and make it difficult to imagine adding another emotionally charged responsibility like abortion care.

> *“There’s such idealism when you’re starting out—thinking birth is always lovely, always about mothers and babies starting life together. And of course, the reality is that it’s not. There’s frankly, so many things that can go wrong, and so many sad and poor outcomes. I can imagine that people who have worked on labor and delivery for a while could feel differently, maybe hardened by their jobs . . . they may feel less positive about abortion.”* (1001, midwife)

Others reflected on how working in an under-resourced safety-net hospital with high patient complexity can foster resentment and moral exhaustion:

> *“It’s a safety net hospital, and it’s publicly insured. There’s just a lot of judgments rolling around all the time. I think, without a conscious effort to stay open you can get bitter. I’ll use substance use disorder as an example. I see a lot of people who, somebody comes in, they have a substance, use disorder, they have a pregnancy complication and a bad outcome. And the staff are angry with them. They take it very personally, like they’re very angry with the patient. . . It’s very easy to become very judgmental and very angry over time when there are decisions that are made that you feel are unjust or not right. I think over time, maybe people could relate that to abortion services as well.”* (1014, midwife)

Similar sentiments emerged among NICU staff; one nurse described how years of witnessing infants born into adversity reshaped her views on motherhood and the morality of abortion care:

> *“I approach it from a different place because I’m seeing all these kids that are brought into the world from sex trafficking and from women who don’t know about or are too high to even think about birth control. I’m dealing with babies that aren’t getting the best start in the world. [Working in the NICU] may have shaped how I feel about abortion now compared to how I did 10 years ago.”* (5022, NICU RN)

#### (b) Personal abortion experiences influenced beliefs

The workshop led many participants to reexamine their own abortion experiences, surfacing complex emotions that shaped how they engaged with the workshop and their work; as one nurse described, “*abortion is a really personal decision, and beliefs come from your own experiences”* (6008, NICU RN). For several participants, the session surfaced memories of colleagues and patients whose abortion experiences had left an emotional imprint. Hearing a colleague’s grief after a D&E reshaped one labor and delivery nurse’s understanding of the importance of offering induction abortion on labor and delivery:

> *“I have a coworker that I was telling, “You know that we’re gonna start this [induction abortion] service.” And she was so happy. She said, “I never got to see my 21-week infant for 20 weeks or so, and they did a D&E, so my baby was torn from limb to limb instead of me being able to hold and let my other kids hold the infant and to feed it. We could have taken pictures.” She was really excited that this [induction abortion on L&D] is gonna be offered for women in the future. It happened to her years ago, but she was just saying how wonderful it was. I guess when we had the workshop I thought about her a lot. And how it would have made a difference in her life to have that experience of being able to go to labor, to choose which way she wanted to go.”* (8012, L&D RN)

For others, the workshop helped participants process their own abortions; they acknowledged how these experiences shaped their moral views and fostered compassion for both others and themselves:

> *I think for me, I’m more conservative when it comes to abortion because I’ve had abortions myself. So I kinda just feel that, if we could prevent them, it’s better than actually having to perform them, right? . . . I had two abortions—one when I was still in high school, and another during a complicated time in my life. The second one brought a lot of guilt, anxiety, and depression. It made me feel like abortion was selfish, and I became very anti-abortion. But during the workshop, I heard stories that opened my mind—about people with very sick babies or whose lives were at risk. That helped shift my perspective and understand that there are serious reasons why someone might need this care. And also deal with some of the trauma that I’ve had to experience around it (quote was lightly edited for confidentiality)* (9001, postpartum staff)

#### (c) Post-Roe context heightened VCAT relevance

The workshop was conducted just over a year after *Dobbs v. Jackson Women’s Health Organization* ended the federal guarantee to abortion access. Participants described how abortion “*never really came up*” before *Dobbs* but felt a heightened obligation in California—where access was preserved—to “*meet a need for people*” (3009, NICU RN). Others wondered whether the *Dobbs* decision was the impetus for the workshop; as one NICU nurse reflected, “*I’m furious the government is taking choice away from women and making workshops like this even necessary… Is that why this workshop came to be?*” (5022, NICU RN).

### (4) Respect for autonomy and commitment to patient care emerged as core, shared values

While abortion beliefs varied among participants, all described prioritizing patient autonomy. Even participants who reported neutral or ambivalent beliefs about abortion articulated a strong commitment to respecting patients’ decision-making over their bodies:

> *“I’m kind of like on the fence. I’m not super pro-abortion, I can’t say I’m really against them, either. I don’t think that government should dictate what people’s choices are, that people should be free to choose what they want. So what does that make me? Respectful of other people’s autonomy, I think.”* (6001, L&D RN)

Providers consistently emphasized that personal beliefs “*took a back seat*” to patient needs (3009, NICU RN), and that this shared commitment was *“reassuring”* (3008, Postpartum RN).

> *“Even though we all have different feelings about abortion, that we all agree that a woman’s bodily autonomy is of the utmost importance, that was really powerful to me. [It] made me be sort of more in love with my group and I’m so happy that we all feel this way.”* (1001, midwife)

In addition to prioritizing *patient* autonomy, participants also discussed the importance of *provider* autonomy and that no one “*should be required to do something against their religion or beliefs.”* (6001, L&D RN). When provider and patient autonomy were at odds, participants weren’t always sure how to “*draw the line between personal beliefs and someone else’s needs”* (3004, L&D RN).

## DISCUSSION

In this qualitative study of perinatal care providers preparing to implement induction abortion services on labor and delivery, we found that a VCAT workshop created a structured space for participants to reflect on their values, understand colleagues’ perspectives, and navigate professional disagreement. These findings align with prior work demonstrating that VCAT workshops can increase empathy and support for abortion care among OB-GYN residents and international healthcare workers,^22, 23, 26^ and extend this literature in several important ways. First, we examined VCAT as a required component of abortion service expansion within a multidisciplinary perinatal workforce. Second, by applying an explicit empathy framework, we demonstrate how VCAT supported key components of clinical empathy—cognitive perspective-taking, communication across difference, and, for some participants, intentions to act in ways that support a diverse workforce and broaden the mission of labor and delivery. In a polarized reproductive health landscape, VCAT may be a valuable tool for fostering professional cohesion and acceptability of abortion care.

The strengths of this study include a rich and robust data set, inclusion of multidisciplinary perinatal healthcare providers, and high compliance with workshop participation, as attendance was mandatory. This study also has limitations. It was conducted at a single, urban, hospital in a safe with strong legal protections for abortion, which may limit transferability to other settings, particularly in states with more restrictive abortion legislation. Participants self-selected into interviews after a mandatory workshop, so views may over-represent those who were willing to discuss abortion and under-represent more strongly opposed perspectives. As with all qualitative work, our findings are not intended to be statistically generalizable, rather, they provide an in-depth description of how one multidisciplinary perinatal team experienced VCAT in the context of abortion service expansion.

## CONCLUSIONS AND IMPLICATIONS

VCAT workshops provided a structured, safe opportunity for L&D staff to reflect on the complex personal and professional dimensions of abortion care. The process helped participants explore value tensions, clarify their professional roles, and identify shared ethical commitments to patient care. These findings suggest VCAT may be a valuable tool for preparing multidisciplinary perinatal teams to navigate institutional abortion care expansion with empathy, professionalism and cohesion.

## Data Availability

All data produced in the present study are available upon reasonable request to the authors

## Notes

**Conflicts of interest:** The authors report no conflicts of interest.

### Competing Interest Statement

The authors have declared no competing interest.

### Funding Statement

This study did not receive any external funding.

